# Abnormal renal function tests at presentation in severe COVID 19 pneumonia and its effect on clinical outcomes

**DOI:** 10.1101/2022.10.21.22281382

**Authors:** Mehak Hanif, Kamran Khan Sumalani, Vishal Mandhan, Zarkesh Shaikh, Shahbaz Haider

## Abstract

**Aim:** To determine the incidence of abnormal renal function tests at presentation in South Asian patients admitted with severe COVID 19 pneumonia and determine its effect on disease severity and clinical outcomes

**Methods:** This was a retrospective cross-sectional study conducted at the COVID Intensive care unit of a large tertiary care government hospital in Karachi, Pakistan. 190 patients admitted over five months from 1/5/2021 till 30/6/2021 were included in the study. Patient demographic characteristics, comorbidities, and clinical manifestations of COVID 19 infection were recorded. Laboratory values at the time of presentation, including Hemoglobin, NLR, platelets, blood urea nitrogen, glomerular filtration rate (GFR), inflammatory markers, liver function tests, and electrolytes were recorded. Patient outcome and need for mechanical ventilation were assessed 28 days after admission and compared with the incidence of abnormal renal functions at presentation.

**Results:** Mean GFR and BUN at presentation were 69.7 and 28.4 respectively. 109 (50.4%) patients had abnormal renal function tests at the time of presentation. 76 (40.0%) patients had low GFR and 33 (17.4%) had only raised BUN with normal GFR. Mean GFR was lower in non-survivors vs survivors (p-value 0.000) and in patients who required mechanical ventilation (p-value 0.008). Patients who had low GFR showed greater mortality than those with normal GFR (p-value 0.04) and were more likely to require mechanical ventilation (p-value 0.04).

**Conclusion:** Low GFR at presentation is common in patients with severe COVID 19 pneumonia and is associated with a higher in-hospital mortality rate and need for mechanical ventilation.

## INTRODUCTION

The spectrum of SARS Cov 2 infection ranges from asymptomatic infections to symptomatic disease with mild-severe symptoms ^(1)^. Severe illness is characterized by SpO2 <94% on room air at sea level, a ratio of arterial partial pressure of oxygen to fraction of inspired oxygen (PaO2/FiO2) <300 mm Hg, respiratory frequency >30 breaths/min, or lung infiltrates >50% on imaging ^(2)^. Among hospitalized patients, the proportion of critical or fatal diseases is higher ^(3)^. Among those who are critically ill, respiratory failure from Acute respiratory distress syndrome (ARDS) is the dominant finding ^(4)^.

Studies have shown that patients with COVID 19 can have abnormal kidney function tests and evidence of acute kidney injury during hospitalization ^(5)^. The ability of the virus to target angiotensin-converting enzyme (ACE) 2 receptors, allows it to target several organs, including the lungs and the kidneys ^(6)^. The pathophysiology and mechanisms of Acute kidney in COVID 19 patients are not fully understood. Kidney involvement can range from the presence of hematuria and proteinuria to acute kidney injury requiring renal replacement therapy ^(7)^. Renal complications are associated with higher mortality in COVID 19 patients ^(8)^.

Previous studies were done in China report an incidence of acute kidney injury from 0-15% in hospitalized patients. However, studies from the US report a higher incidence of acute kidney injury from 14-69% amongst hospitalized patients ^(9)^. We carried out our study in the Intensive care unit of one of Pakistan’s largest tertiary care public hospitals, Jinnah postgraduate medical center. We aimed to determine the incidence of abnormal renal function tests in patients with severe SARS Cov2 pneumonia and determine its effect in predicting disease severity and clinical outcomes.

### OBJECTIVE

To determine the frequency of abnormal renal function tests at presentation in patients admitted with severe COVID 19 pneumonia in a tertiary care hospital and determine its effect in predicting disease severity and clinical outcomes

## MATERIALS AND METHODS

This was a cross-sectional study conducted at the COVID isolation ward of Jinnah postgraduate medical center, one of the largest tertiary care government hospitals in Karachi, Pakistan. Patients admitted over a period of five months from 1/5/2021 till 30/6/2021 were included in the study. Total number of SARS cov2 positive patients admitted during the period of study was 372. After application of inclusion and exclusion criteria, we were able to obtain the data of 190 patients. The study was approved from the ethical committee of Jinnah postgraduate medical center. Inclusion criteria included adult patients aged >18years and positive PCR test for SARS cov2. Exclusion criteria included previous respiratory pathologies including interstitial lung disease, chronic pulmonary disease and pulmonary tuberculosis; heart failure; chronic liver disease or chronic kidney disease; history of malignancy; use of immunosuppressive drugs including long term steroids; sepsis; recent major surgical procedures; active urinary tract infection or urinary tract obstruction; suspicion of drug-induced renal injury and hypertensive crisis.

Glomerular filtration rate (GFR) was calculated using the CKD-EPI equation ^(10)^.

GFR = 141 * min (Scr/κ,1)^α^ * max(Scr/κ, 1)^−1.209^ * 0.993^Age^ * 1.018 [if female] * 1.159 [if black]. Creatinine clearance (CrCl) was calculated using the Cockcroft-Gault equation ^(11)^.

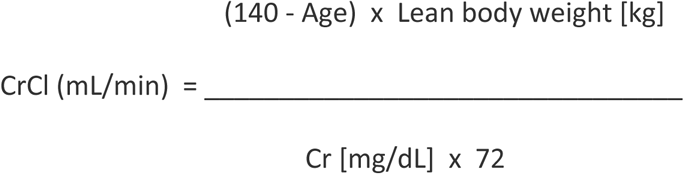

For women, the formula requires multiplication by 0.85 to account for smaller muscle mass compared with men.

Abnormal renal function tests were defined as Glomerular filtration rate <60 ml/min, based on the normal values suggested by National Kidney Foundation and Kidney Disease Improving Global Outcome (KDIGO) or blood urea nitrogen (BUN) less than 24mg/dl, which was the normal limit of the laboratory of our hospital ^(12) (13)^.

After informed consent, blood samples of all patients are drawn for routine laboratory tests within 24 hours of admission. The routine tests include arterial blood gas analysis, complete blood count, liver function tests, renal function tests, coagulation profile and inflammatory markers (C reactive protein, Lactate dehydrogenase, D Dimers and Ferritin). Patients are also sent for Chest X-rays within 24 hours of admission to determine the degree of lung involvement. This data collected within 24 hours of admission was used during the study. Other demographic characteristics of patients; including age, gender and comorbidities were also recorded. The variables of outcome were recorded by documenting the total duration of hospital stay, total duration of oxygen therapy, total duration of invasive or non-invasive ventilation and clinical status of the patient at 28 days post admission.

Data was analyzed using SPSS version 26. Mean and Standard deviation was calculated for quantitative variables like Hemoglobin, Total leukocyte count, Neutrophil lymphocyte ratio, Platelets, Urea, Creatinine, Glomerular filtration rate, CRP, LDH, and D-Dimers. Frequencies and percentages were used for qualitative data, including gender and comorbidities. The rest of the data was organized into categorical variables, including PiO2/FiO2 ratio, oxygen requirement, duration of hospital stay, duration of oxygen therapy, and duration of invasive and non-invasive ventilation. The Chi-Square test was used for the analysis of categorical data. An Independent Sample t-test was used to check inter-categorical differences of mean values. P values of less than 0.05 were considered statistically significant.

## RESULTS

Amongst those studied, 62.1% were males and 37.9% were females. The mean age of the patients was 57 years. 50.7% of patients had diabetes mellitus, 44.7% of patients had hypertension and 6.8% had ischemic heart disease. Most patients (51.6%) had PO_2_/FiO_2_ ratio less than 100 at presentation. Patient demographics, comorbidities and clinical manifestations are summarized in Table 1.

**Table 1:**
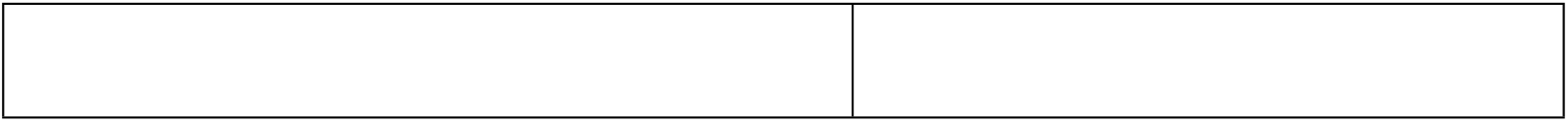
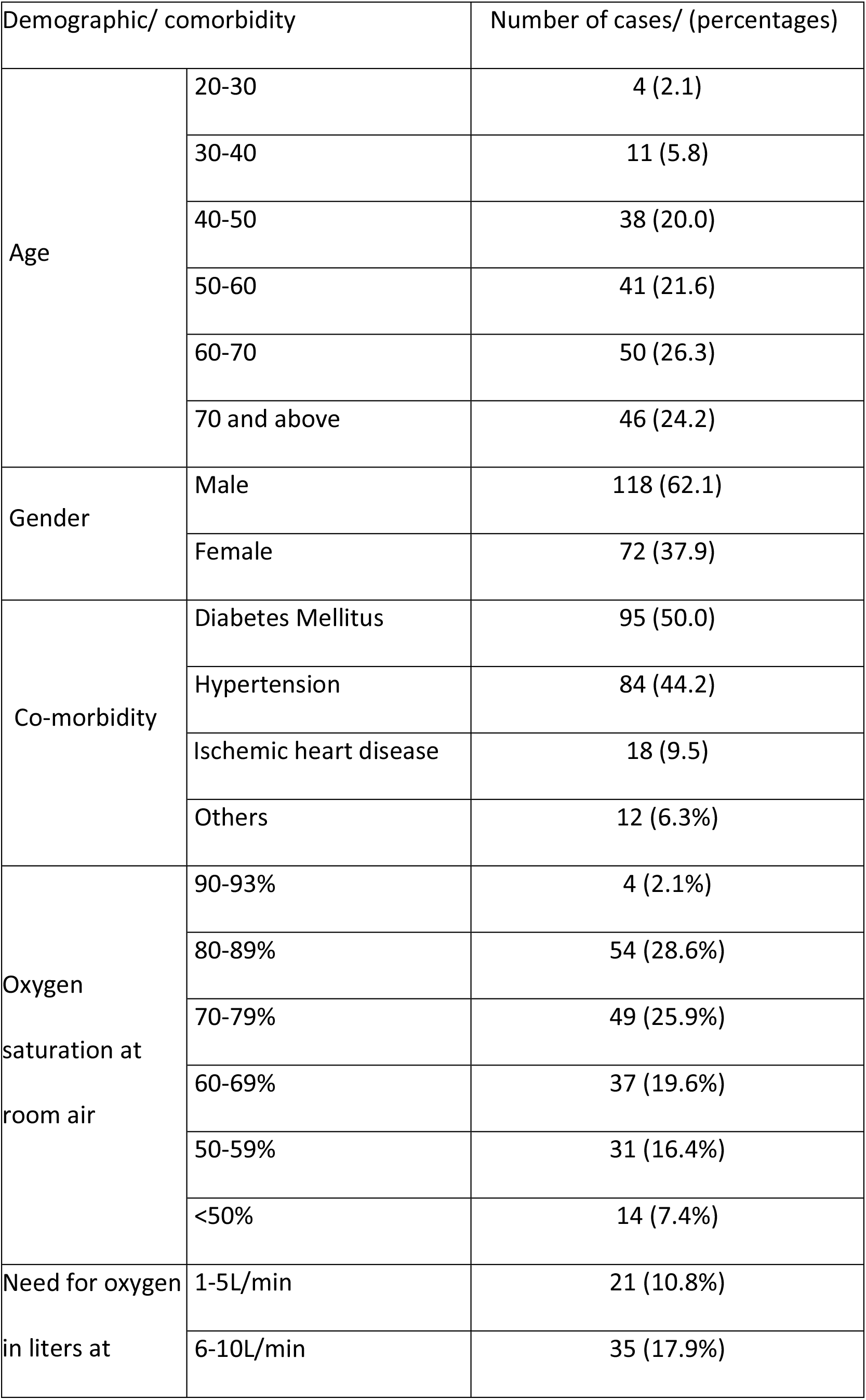
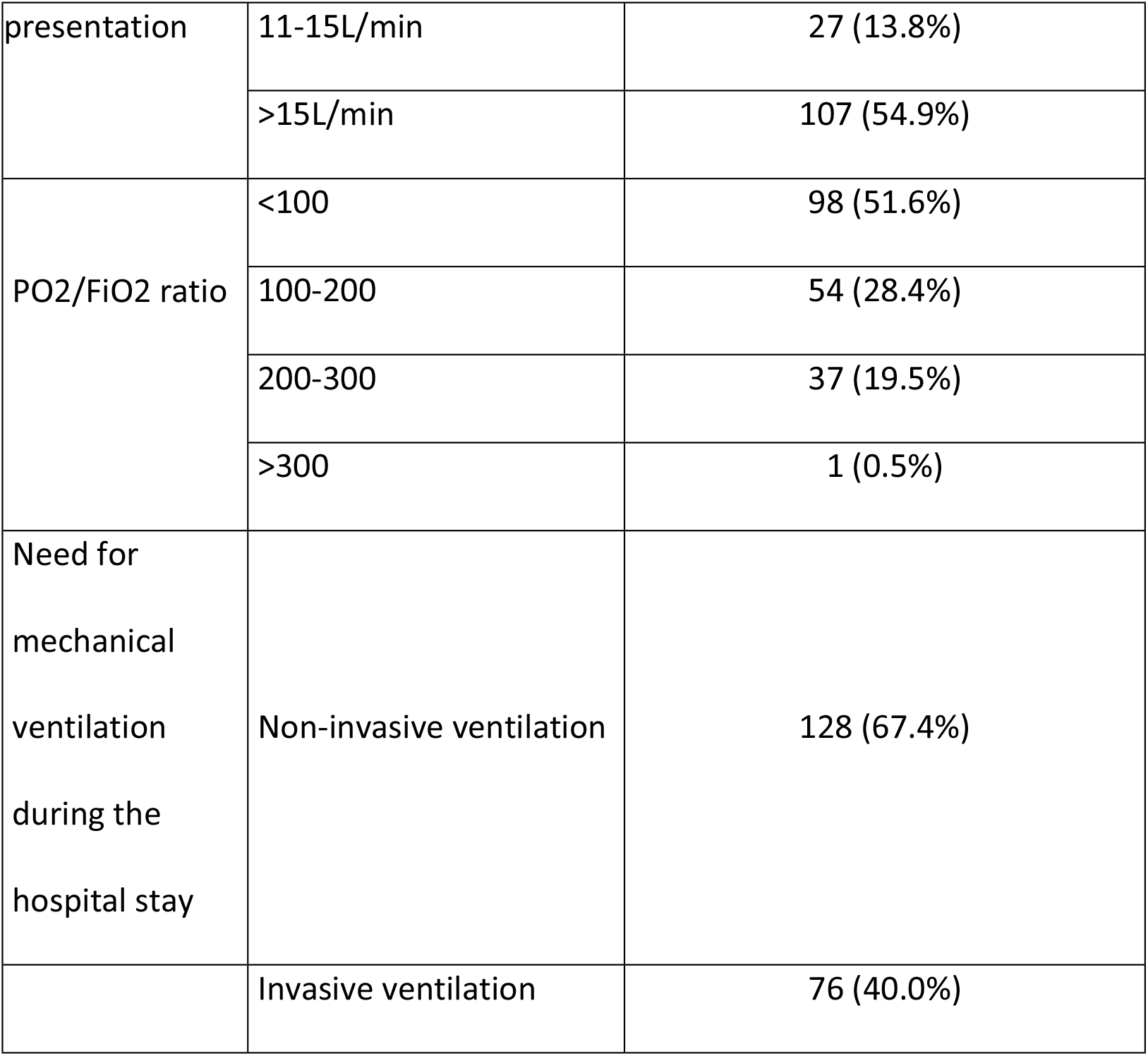
Patient demographics, comorbidities, and clinical characteristics

Mean BUN at presentation was 28.4 mg/dl, CrCl 75.9 ml/min and GFR 69.7 ml/min. 109 (50.4%) patients had abnormal renal function tests at the time of presentation. 76 (40.0%) patients had low GFR and 33 (17.4%) had only raised BUN with normal GFR. The laboratory characteristics of patients at presentation are summarized in Table 2.

**Table 2:** Laboratory characteristics at presentation

| Laboratory value at presentation | Mean +/- Standard deviation |
| --- | --- |
| Hemoglobin | 12.4± 1.9 |
| Total leukocyte count | 11.9 ± 5.0 |
| Neutrophil lymphocyte ratio | 11.1 ± 8.2 |
| Platelets | 238.5±99.9 |
| Blood Urea Nitrogen | 29.1±19.8 |
| Creatinine | 1.4±1.5 |
| Glomerular filtration rate | 69.7±30.6 |
| C Reactive Protein | 126.6±148.7 |
| D Dimers | 4.4±5.9 |
| Lactate Dehydrogenase | 825.3±486.8 |
| Ferritin | 903.1±573.9 |
| International Normalized ratio | 1.1±0.8 |
| Serum sodium | 137.4±6.0 |
| Serum potassium | 3.9±0.7 |
| Total bilirubin | 0.6±0.4 |
| Alkaline phosphatase | 150.5±110.4 |
| Alanine aminotransferase | 65.9±138.4 |

All patients received bolus doses of IV methylprednisolone followed by dexamethasone. 74.6% of patients received Remdesivir. Only 4.2% of patients received Tocilizumab due to non-availability. 73.0% patients received treatment dose anticoagulation (enoxaparin/heparin) while 26.5% received prophylactic dose anticoagulation. Two patients didn’t receive anticoagulation because of contraindications. 58.2% of patients with a high cardiovascular disease risk score were also given aspirin.

The outcome variables of patients included in the study are summarized in Table 3. Most patients stayed in the hospital for 0-7 days (38.1%) and 7-14 days (34.9%). 6.3% of patients stayed for 28 days. Most patients received oxygen therapy for 0-7 days (40.5%) and 7-14 days (34.2%). 6.8% of patients required oxygen therapy for more than 28 days. 65.3% of patients required non-invasive ventilation. Out of those who received non-invasive ventilation, most patients (44.7%) received it for 0-7 days. 2 patients (1.1%) required it for more than 28 days. 41.6% of patients required invasive ventilation. Out of those who received invasive ventilation, 81.4% of patients received it for 0-7 days, only 2 patients (2.4%) received it for more than 14 days.

**Table 3:** Outcome

| Outcome after<br>28 days | Outcome | Number of cases/(percentages) |
| --- | --- | --- |
|  | Death | 117 (61.6%) |
|  | Discharged on room air | 54 (28.4%) |
|  | Discharged on home oxygen | 4 (2.1%) |
|  | Admitted with oxygen support via mask/nasal cannula | 186 (97.9%) |
|  | Admitted with non-invasive ventilation | 7 (3.7%) |
|  | Admitted with invasive ventilation | 1 (0.5%) |
| Duration of hospital stay | 0-7 days | 73(38.4%) |
|  | More than 7 days | 117 (61.6%) |
| Duration of oxygen therapy | 0-7 days | 76 (40%) |
|  | More than 7 days | 114 (60%) |
| Duration of non-invasive ventilation | 0-7 days | 84 (44.2%) |
|  | More than 7 days | 44 (23.1%) |
| Duration of invasive ventilation | 0-7 days | 68 (35.8%) |
|  | More than 7 days | 8 (4.2%) |

23 (12.6%) patients required dialysis. All patients (100%) who required dialysis were on ventilators.

By 28 days post-admission, most patients (61.6%) died. 28.4% were discharged on room air, 4.2% remained admitted with oxygen support, 3.7%remained admitted with non-invasive ventilation and 2.1% were discharged on home oxygen.

Tables 4 and 5 compare the incidence of low GFRand mean GFR in different groups of patients respectively. Abnormal renal function tests did not show any significant association with patient comorbidities. We did not find any significant difference in lab parameters including neutrophil-lymphocyte ratio, platelets, alanine aminotransferase, and C-reactive protein in patients with low GFR vs those with normal GFR. Patients who were more than 60 years old were more likely to present with low GFR than younger patients (p-value 0.001). Mean GFR was significantly lower in non-survivors vs survivors (p-value 0.001) and in patients who required mechanical ventilation (p-value 0.008). Patients who had low GFR showed greater mortality than those with normal GFR (p-value 0.04) and were more likely to require mechanical ventilation (p-value 0.04). However; when compared with patients with normal GFR, patients with low GFR did not show any significant variance with the duration of hospital stay, oxygen therapy, non-invasive ventilation, and invasive ventilation.

**Table 4:**
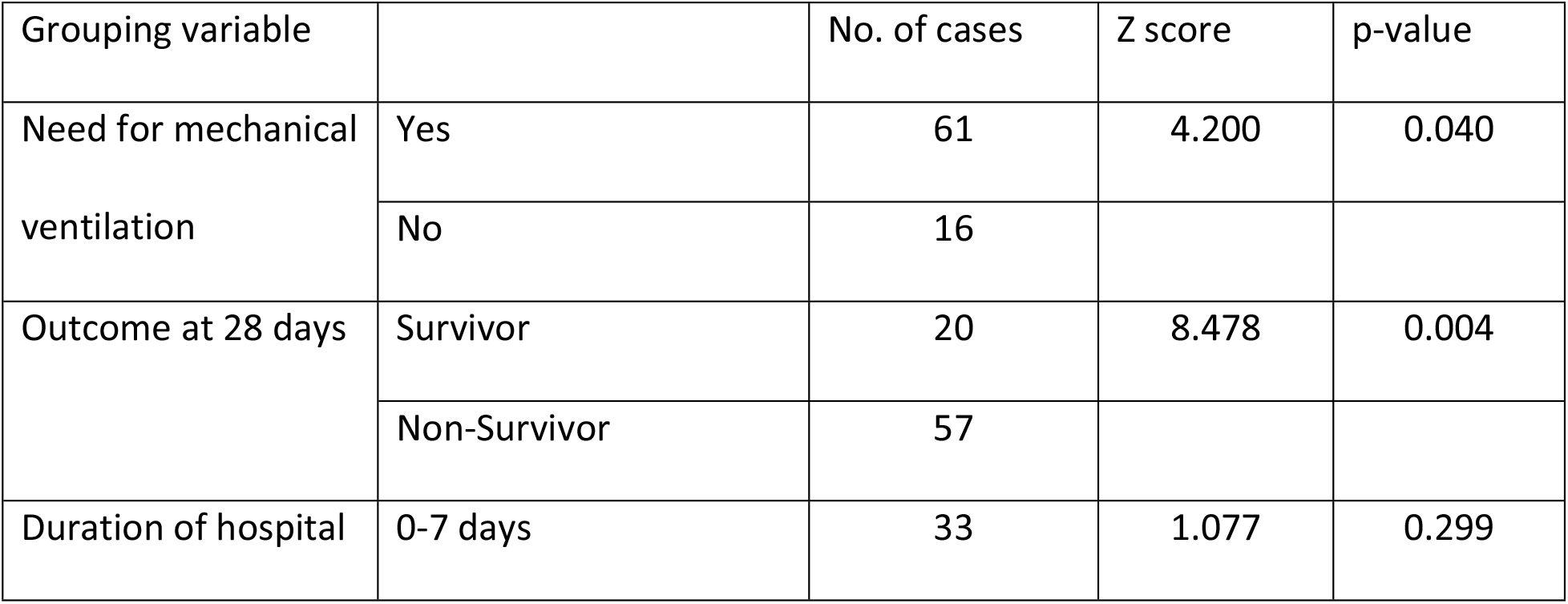
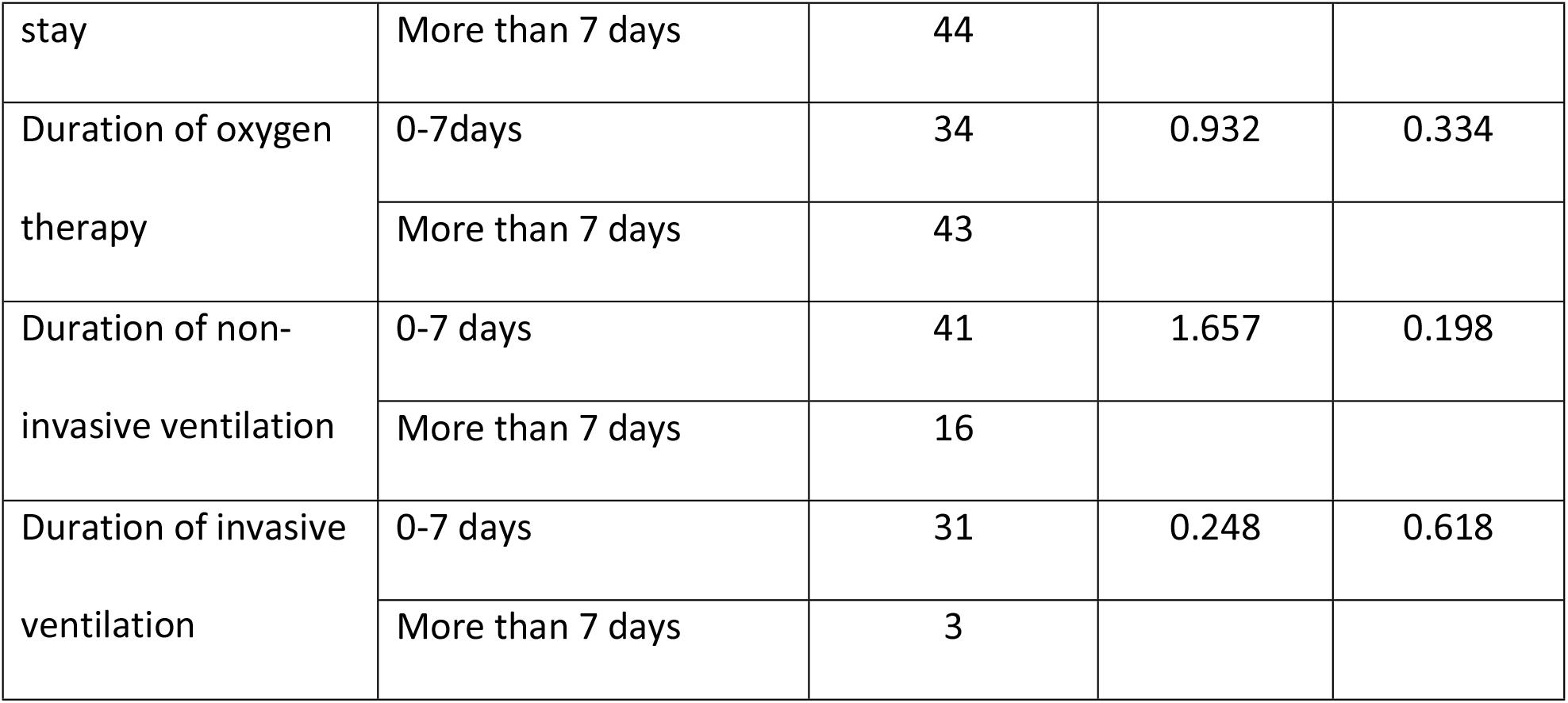
Chi-square test – Comparison of the incidence of low GFR in different groups of patients

**Table 5:**
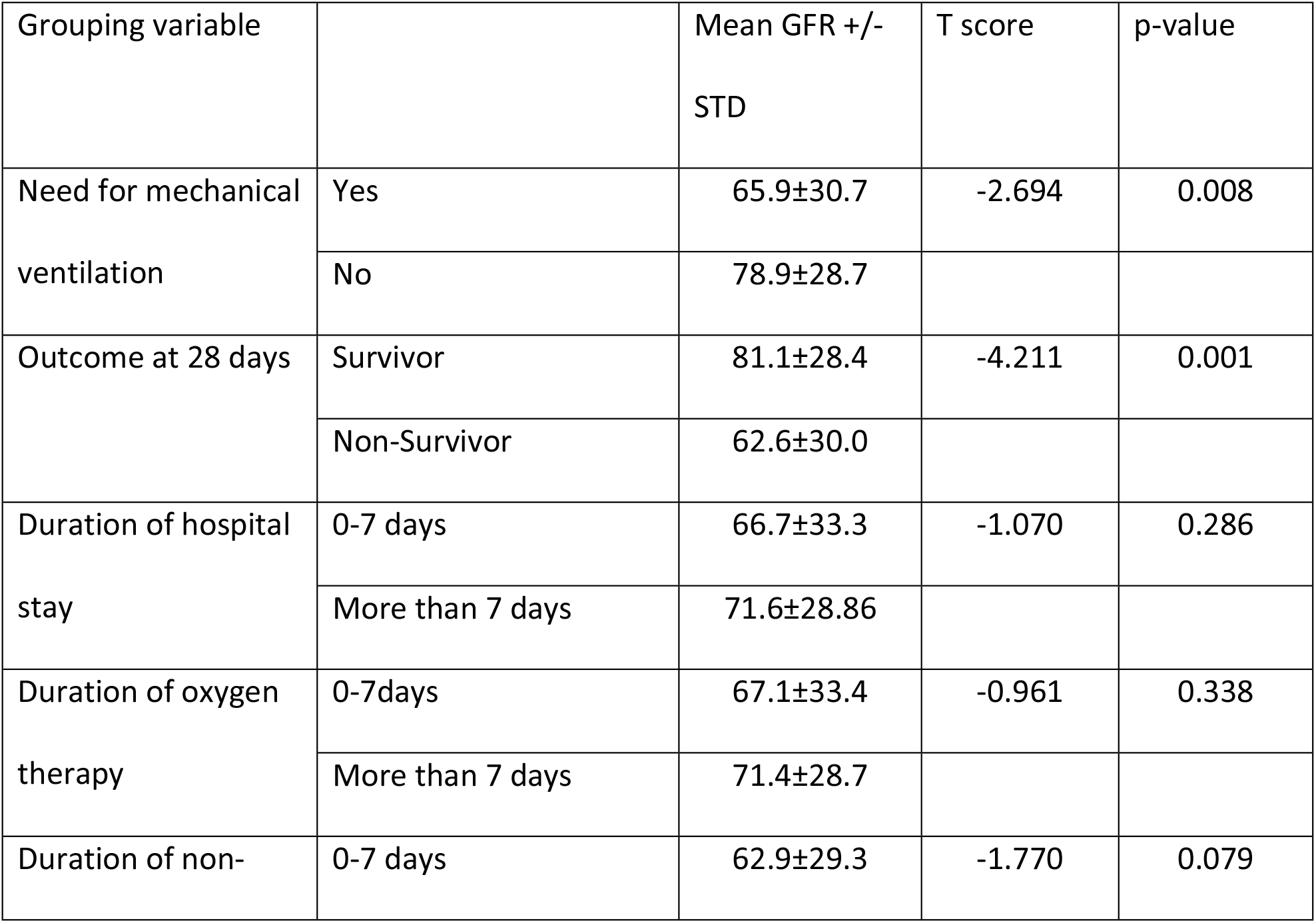
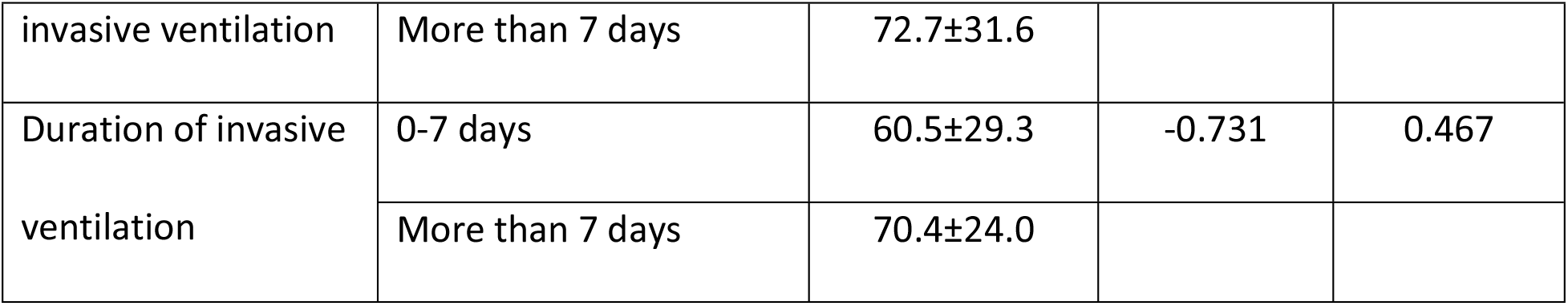
Independent sample test – Comparison of mean GFR in different groups of patients

## DISCUSSION

We studied the incidence of abnormal renal function tests in patients with severe COVID 19 pneumonia and determined its effect on clinical outcomes and disease progression. Many studies, to date, have recorded the incidence of abnormal renal function tests in mild and severe COVID 19 cases. Some studies have also compared the predictive effect of GFR on mortality. However, there is a lack of large cohort studies in the South Asian population. Our study was carried in one of the largest, yet resource-limited, public sector tertiary care hospitals in Pakistan.

The mechanism of renal injury in patients with COVID-19 is complex and multifactorial. Firstly, SARS Cov 2 PCR fragments have been discovered in the blood and urine of COVID 19 patients and it has been hypothesized that the virus exerts direct cytopathic effects on the kidneys ^(6)^. SARS Cov 2 uses angiotensin-converting enzyme 2 (ACE2) as a cell entry receptor and expression of the receptor in the kidney is very high ^(14) (15)^. Thirdly, virus-induced cytokine release might cause direct renal injury, or indirectly affect the kidney because of hypoxia, shock, or rhabdomyolysis ^(16–19)^.

In our study 17.4% of patients had an isolated increase in BUN, with normal GFR. The most common cause of raised BUN is a renal disease leading to reduced clearance of urea from the blood. However, the limitation of urea as a test of renal function is that plasma urea is not a sufficiently accurate reflection of reduced GFR. Urea may be raised despite a normal GFR, so BUN is not a specific test for renal function ^(20)^. Extrarenal causes of raised urea in hospitalized or critically ill patients include dehydration, heart failure, trauma, severe infection, starvation, and use of corticosteroids, which could have accounted for the raised BUN and normal GFR in our patients ^(21)^.

Multiorgan involvement has been seen in patients with COVID 19 ^(22)^ and studies have shown lower platelet and lymphocyte counts, higher leukocyte count, and a higher rate of comorbidities in COVID 19 patients who developed acute kidney injury ^(6)^. However, our study did not show any significant difference in comorbidities or lab parameters in patients with abnormal renal function tests vs those with normal tests.

In our study, we excluded patients who had known chronic kidney disease, so that we could have an estimate of the incidence of acute kidney injury in patients with COVID 19. The incidence of abnormal renal functions at presentation was 57.4%, out of which 40.4% of patients had low GFR and 17.4% had high BUN with normal GFR. 12.6% of patients required dialysis. Many other studies have also shown that COVID 19 is associated with Acute Kidney injury or abnormal renal functions at presentation. A study done on a smaller cohort of patients in Pakistan, earlier in the course of the epidemic, showed that 39.4% of patients had raised creatinine levels at the time of presentation ^(23)^. Another study done on 193 patients in China, early in the course of the epidemic showed that 31% of patients had an elevated level of blood urea nitrogen (BUN) and 22% had increased serum creatinine ^(24)^. In one observational study, carried out at the same time, on 5,449 hospitalized patients in New York City, the incidence of AKI was 36.6% with 14.3% of patients requiring dialysis (30%) ^(25)^.

Our study showed a higher incidence of abnormal renal function tests than these studies. Firstly, this could be because our study was done at a public hospital in Pakistan. Pakistan is a country that struggles with low literacy and health awareness and patients usually present later in the course of the disease ^(26)^. Secondly, our study included only cases with severe COVID 19 pneumonia and not mild cases. Thirdly, our study was done later in the course of the epidemic, when more aggressive strains of COVID 19 had emerged ^(27) (28)^.

Our study revealed that patients with low GFR at the time of presentation had higher mortality and need for mechanical ventilation than those with normal GFR. However, there were no significant differences in duration of hospital stay, oxygen therapy, invasive or non-invasive ventilation in the two groups. Several studies have shown that low GFR at the time of presentation is associated with a greater number of complications and in-hospital mortality in COVID 19 patients ^(29–31)^.

The main limitations of our study were the wide clinical heterogeneity at presentation. Due to social taboos and lack of awareness, patients seek help at local inexperienced centers and present late in the course of the disease at tertiary care government hospitals in Pakistan.

Additionally, IL-6 inhibitors were not easily available at the hospital. Poor infection control practices were practiced at the hospital because of a lack of logistics and staff awareness. There was a lack of experienced anesthetists at the center. All of these could have had an impact on the clinical course and outcome of patients. Additionally, baseline creatinine of the patients was not available, because of which we could not make an accurate estimation of acute kidney injury in our patients.

In summary, we concluded that abnormal renal function tests at presentation are very common in patients with severe COVID 19 pneumonia, and are associated with a higher in-hospital mortality rate and need for mechanical ventilation. However, larger multicentered trials are needed to further validate the results of our study.

## CONCLUSION

Abnormal renal function tests at presentation are common in patients with severe COVID 19 pneumonia and are associated with a higher in-hospital mortality rate and need for mechanical ventilation. On admission, eGFR can be used as a prognostic marker for mortality and a tool for risk stratification in COVID 19 patients.

## Data Availability

All relevant data are within the manuscript and its Supporting Information files.

